# COLOFIT: Development and internal-external validation of models using age, sex, faecal immunochemical and blood tests to optimise diagnosis of colorectal cancer in symptomatic patients

**DOI:** 10.1101/2024.03.01.24303196

**Authors:** CJ Crooks, J West, J Jones, W Hamilton, SER Bailey, G Abel, A Banerjea, CJ Rees, A Tamm, BD Nicholson, SC Benton, N Hunt, COLOFIT Research Group, DJ Humes

## Abstract

**Background:** Colorectal cancer (CRC) is the 3^rd^ most common cancer in the United Kingdom and is the 2^nd^ largest cause of cancer death.

**Aim:** To develop and validate a model using available information at the time of Faecal Immunochemical testing (FIT) in primary care to improve selection of symptomatic patients for CRC investigations.

**Methods:** All adults ≥ 18 years of age referred to Nottingham University Hospitals NHS Trust between 2018 and 2022 with symptoms of suspected CRC who had a FIT. Predicted 1-year CRC diagnosis were calculated, and externally validated, using Cox proportional hazards modelling with selected multiple fractional polynomial transformations for age, faecal haemoglobin concentration (f-Hb) value, mean corpuscular volume (MCV), platelet count and sex.

**Results:** At a CRC risk threshold of 0.6% (equivalent to f-Hb=10 µgHb/g (µg/g)) overall performance of the validated model across age strata using Harrell’s C index was ≥ 0.91% (overall C-statistic 93%, 95% CI 92%-95%) with acceptable calibration. Using this model yields similar numbers of detected and missed cancers but require ∼20% fewer investigations than a f-Hb ≥10 µg/g strategy. For approximately 100,000 people per year with symptoms of suspected CRC, we predict it might save >4,500 colonoscopies with no evidence that more cancers would be missed if we used our model compared to using FIT f-Hb≥10 µg/g.

**Conclusions:** Including age, sex, MCV, platelets and f-Hb in a survival analysis model to predict the risk of CRC yields greater diagnostic utility than a simple binary cut off f-Hb≥10 µg/g.

**Transparency statement:** The lead author and manuscript’s guarantor (CJC) affirms that the manuscript is an honest, accurate and transparent account of the study being reported; than no important aspects of the study have been omitted: and that any discrepancies from the study as originally planned have been explained.

**Role of the funding source:** This project was funded by the National Institute for Health and Care Research (NIHR) [Health Technology Assessment (HTA) Programme (Project number 133852); awarded to CJR, WH & LS] and will be published in full in the HTA journal. Further information is available at: [https://fundingawards.nihr.ac.uk/award/NIHR133852]. The views expressed are those of the authors and not necessarily those of the NIHR or Department of Health and Social Care and sponsored by Nottingham University Hospitals NHS Trust. The funder and sponsor had no role in the study design, in the collection, analysis, and interpretation of data; in the writing of the report; and in the decision to submit the article for publication. We confirm the independence of researchers from funders and that all authors, external and internal, had full access to all of the statistical reports and tables in the study and can take responsibility for the integrity of the data and the accuracy of the data analysis. SERB was supported by an NIHR Advanced Fellowship while undertaking this work (NIHR301666) and received additional support from the Higgins family. BDN was supported by a National Institute of Health Research Academic Clinical Lectureship and a CRUK Research Careers Committee Postdoctoral Fellowship (RCCPDF\100005).

**Ethics approval statement:** HRA and Health and Care Research Wales (HCRW) approval was given for this study - IRAS project ID: 312362; Protocol number: 22ON007; REC reference: 22/HRA/2125; Sponsor: Nottingham University Hospitals NHS Trust.

## Introduction

Colorectal cancer (CRC) is the 3rd most common cancer in the United Kingdom and is the 2^nd^ largest cause of cancer death^1^. Bowel cancer screening programmes reduce mortality but takes a long time to have an effect. Currently, screening only accounts for approximately 10% of CRC diagnoses – the majority occur through symptomatic patients being referred with a suspicion of cancer through a variety of pathways^2^ ^3^. Additionally, 20% of CRCs present as an emergency^4^. The “risk threshold” for urgent referrals for investigation was set at a 3% positive predictive value of cancer by the English National Institute for Health and Care Excellence (NICE), i.e. those referred by primary care on a cancer diagnostic pathway should have a risk of a specific cancer of 3% or more^5^. Faecal Immunochemical Testing for haemoglobin (Hb) (FIT) identifies haemoglobin in faeces as an indicator of possible CRC. This is the approach used in the English asymptomatic population-based bowel cancer screening programme (at a higher threshold than used in symptomatic patients). FIT for symptomatic patients was recommended by NICE in 2023 with a FIT cut-off of ≥10 µgHb/g (µg/g) set for this purpose^6^. However, current demand for colonoscopy or computed tomography colonography (CT-colonography) capacity in the UK and many other countries far outstrips capcity^7^ ^8^. This imbalance between demand for investigation at f-Hb ≥10 µg/g and supply of colonoscopy or CT-colonography means that investigations will be delayed for some people at higher risk of CRC whilst many normal investigations are being performed. This situation has been worsened by the COVID-19 pandemic^9^.

Recent evidence from Nottingham suggests that stratifying by age and the presence or absence of anaemia could identify those people with a FIT ≥10 µg/g at a low CRC risk, well below the defined 3%, who do not need investigation^10^. While stratified approaches could work, an alternative approach, is to use a clinical prediction model to estimate risk of CRC at an individual level to tailor investigation. Such an approach should maintain diagnostic performance whilst decreasing the burden on diagnostic services by reducing the number of colonoscopies and/or CTC’s performed. Information from the patient i.e. f-Hb level, age, sex and blood indices could be used to inform whom to investigate based on their predicted risk of CRC. Such predictions could inform all stakeholders (patients, general practitioners, policy makers) as to who could either safely avoid investigation or have it routinely (and be reassured that the risk of CRC is low) whilst prioritising those with the highest risks of CRC for urgent colonic investigations. A recent systematic review highlighted the potential merits of this approach using f-Hb; however, it identified only three models with internal and external validation FAST, COLONOFIT and COLONPREDICT but it concluded that models to date had been developed with poor methodology and further work was required to develop clinically relevant models with internal and external validation and a net benefit approach to compare outcomes to current pathways^11^.

Our aim was to develop and validate a clinically useful prediction model to estimate 1-year risk of CRC using all people in the referral population for Nottingham University Hospitals NHS Trust who had completed a FIT in primary care. We planned to compare a model-based strategy to the NICE f-HB cut-off of 10 µg/g and a risk of referral threshold of 3% (f-Hb ≥ 40 µg/g and others including a 1% and 2% risk) akin to the NICE early cancer diagnosis recommendations and assess diagnostic utility using a net benefit approach^6^.

## Methods

### Nottingham Rapid Colorectal Cancer Diagnosis Pathway (NRCCD)

In Nottingham since November 2016, a locally commissioned pathway allows FIT to be requested by general practitioners as a triage tool for all symptomatic patients referred with suspected CRC, except those with rectal bleeding (who were not at this point eligible for a FIT test) or a palpable rectal mass who were not at this point eligible for a FIT test. From November 2021 general practitioners were also able to request the test for those with rectal bleeding. In addition, a Full Blood Count (FBC) blood test was mandated for all CRC referrals irrespective of symptoms or age.

The study is reported consistent with the TRIPOD (transparent reporting of a multivariable prediction model for individual prognosis or diagnosis) guidelines^12^. The study was undertaken as part of the COLOFIT programme of work seeking to establish the optimal role of FIT in the clinical pathway.

### Study setting

The study was conducted at Nottingham University Hospitals (NUH) NHS Trust, using data for all primary care requested FIT results, processed within pathology services at Nottingham University Hospitals NHS Trust (NUH) among^13^:

- Adults (≥ 18 years of age)
- Patients within Nottinghamshire registered at a General Practice that would refer to Nottingham University

Hospitals (Nottingham City and South Nottingham Integrated Care Partnerships)

- From 01/Nov/2017 until 31/Nov/2021 for a derivation cohort.
- From 01/Dec/2021 until 31/Nov/2022 for a validation cohort.

All other patients were excluded. FIT requests and results reporting was electronic. FIT dispatch and return were postal from the laboratory and were required for all referred patients to NUH. The kits were distributed and analysed according to manufacturer’s protocols by our accredited FIT laboratory using an OC-Sensor™ platform (Eiken Chemical Co., Tokyo, Japan) as previously described^14^.

### Data Management

The variables of interest were extracted and linked by patients’ unique identification numbers using Microsoft SQL Server from our Trust’s Enterprise Data Warehouse as previously described^10^. The data were anonymised before being accessed by the researchers, so the researchers had no access to identifiable patient level data: no patient level data left NUH NHS Trust. The anonymous data for analysis were analysed on a secure SQL server within NUH that only the analytical team could access for analysis (CC, JW)^10^.

#### Outcomes

CRC was defined from linked Infoflex (Civica) data where all cancers diagnosed at NUH NHS Trust are recorded. Fact and date of death were obtained from the NHS personal demographics service and underlying cause of death (coded with ICD-10) from https://www.hed.nhs.uk/Info/. Patients were followed up for one year for CRC diagnosis or death.

#### Predictors

Each individual had their first recorded FIT (index FIT) identified and were subsequently linked to all the required datasets within NUH NHS Trust’s Enterprise Data Warehouse as previously described^10^. This included age (at date of FIT) using year of birth, sex (defined as male/female), and recorded ethnicity (categorised as White, Black, Asian, Other and unrecorded). Measures of ferritin, iron, transferrin saturation and faecal calprotectin were extracted. Blood test results included haemoglobin/mean corpuscular volume (MCV)and platelets as thrombocytosis has been shown to be associated with undiagnosed colorectal cancer^15^. Missing blood tests values were assumed to be missing at random and imputed separately in the derivation and validation cohorts using two level multiple imputation-chained equations by predictive mean matching, with a random intercept for each patient and time from test fitted as a within patient gradient. Ten imputed datasets were used with up to 10 iterations per dataset. Adequate mixing of imputed values was assessed visually with plots. Additional predictors in the imputation model included age, sex, CRC, and death. We were not able to disaggregate by sex and gender as only sex is recorded electronically.

### Statistical analysis

#### Predictive model building

We constructed a multivariable regression model using F-HB result, age, sex, and haemoglobin, platelets and MCV test results as potential predictors with the outcome of CRC diagnosis within one year of the FIT. A Cox proportional hazards survival model was selected as the primary model to account for censoring from non-CRC deaths. A multivariable selection algorithm was used to select fractional polynomial transformations for f-Hb, haemoglobin, MCV, platelets and age. This used backward elimination with weighted likelihood ratio testing across the stacked imputed datasets whilst keeping the familywise error rate at p = 0.05^16^. Individual level weights were calculated as the smallest proportion of non-missing data across the imputed values divided by the number of imputed datasets following Morris et al^16^. All pairwise interactions between age, f-Hb, platelets and MCV were tested using generalised likelihood ratio tests incorporating all the transformed components for each predictor^17^.

We also developed a logistic regression model with the binary outcome of CRC at one year ignoring censoring, using the same model selection approach as for the Cox model. This was to assess whether the Cox survival analysis was influenced by a change in time to diagnosis of cancer from the FIT rather than predicting the risk of cancer itself. It is presented in the supplementary material.

All analyses were carried out using R^16^ within R Studio.

#### Model targeted validation

The equation developed in the derivation cohort was applied to the Nottingham validation cohort of patients having a FIT with the imputed blood test values^18^. At the time of analysis we only had access to one year of test results for validation subsequent to the derivation cohort. Therefore, for the following performance measures only first FIT per patient excluding repeat tests in subsequent years was used so that patients were not included in both the validation and derivation samples.

Concordance was then measured using Harrel’s C-statistic across the imputed datasets and pooled using Rubin’s rules. Concordance was stratified by age and ethnicity to assess for potential inequalities in the performance of the model. The calibration of the models was assessed by plotting the observed 1-year Kaplan Meier survival probability against the expected 1-year survival predicted by the fitted model, stratified by the linear predictor in deciles of patients with CRC (to avoid strata with few events). The sensitivity, specificity, true positive, false positive, true negative and false negative values and rates were calculated for different test thresholds for the logistic and survival models in the validation and derivation cohorts. To account for censoring in the survival models we calculated false positive (FPV) and true negative values (TNV) as the observed CRC free survival in patients above and below each selected threshold respectively, using Kaplan Meier estimates similar to the approach described by Vickers et al^19^. The false positive and true positive rates were then calculated by multiplying FPV and (1-FPV) respectively by the proportion of patients above the selected threshold. True and false negative rates were calculated as the difference between the false and true positive rates and the overall observed CRC survival and risk respectively.

These estimates were then used to calculate the net benefit (an overall assessment of the weighted difference between the true positives correctly identified and false positives unnecessarily referred). and extrapolated (by multiplying by 100,000) to indicate the number of potential missed cancers and reduction in colonoscopies per 100,000 FITs. These performance estimates were compared to a f-Hb only model with a threshold of ≥10 µg/g and ≥40 µg/g (f-Hb ≥10 µg/g as a currently recommended threshold and f-Hb ≥ 40 µg/g as equivalent to the 3% risk cut off recommended urgent cancer referrals), and an intermediate model with f-Hb, age and sex including fractional polynomial transformations for f-Hb and age. 95% confidence intervals were calculated with bootstrapping, resampling each of the imputed datasets with replacement (n = 1000) and pooling the subsequent estimates to calculate the 2.5% and 97.5% quantiles^20^.

We initiated an external validation (using data from a different geographical location) as planned in the statistical analysis plan, but initial work showed poor calibration in the external dataset chosen which was deemed to be due to due to pre-analytical (sample collection) and population differences that precluded calibration in the external dataset chosen. Whilst work is ongoing to further understand these differences we have subsequently undertaken an external validation using data from East Lancashire Hospitals NHS Trust (ELHT) using all patients who had FIT requested for symptoms of suspected colorectal cancer in primary care from 2019-2023 with one year follow up. Blood tests were identified using the same definitions as our derivation cohort. All FITs were performed on an OC-Sensor™ platform (Eiken Chemical Co., Tokyo, Japan). The algorithm was used to predict 1-year risk of colorectal cancer and a comparison of colonoscopies undertaken between the 1% predicted threshold and the current NICE f-HB cut-off of 10 µg/g.

#### Data sharing statement

This work uses data that has been provided by patients and collected by the NHS as part of their care and support. Under the Data Protection Impact Assessment approval for this work (DPIA reference: IG0889) we are unable to share the original data outside Nottingham University Hospitals NHS Trust.

#### Ethics approval statement

HRA and Health and Care Research Wales (HCRW) approval was given for this study - IRAS project ID: 312362; Protocol number: 22ON007; REC reference: 22/HRA/2125; Sponsor: Nottingham University Hospitals NHS Trust.

#### Patient and public involvement

PPI involvement in development of the COLOFIT work was extensive with review by several PPI panels and named PPI representation on the grant application.

#### Funding

This project was funded by the National Institute for Health and Care Research (NIHR) [Health Technology Assessment (HTA) Programme (Project number 133852)) and will be published in full in the HTA journal. Further information is available at: [https://fundingawards.nihr.ac.uk/award/NIHR133852]. The views expressed are those of the authors and not necessarily those of the NIHR or Department of Health and Social Care.

## Results

### Study population

For the derivation cohort 34,435 patients had 37,216 FIT results recorded between 01/Nov/2017 and 31/Nov/2021, after limiting FIT results to one per year per patient and excluding 2,558 repeat tests within 12 months. Between 1/Dec/2021 and 31/Nov/2022 there were 21,012 patients; however, after excluding those that were repeat tests within 12 months of a previous test (n= 1948) there were 20,234 FITs recorded for the validation cohort. Summary characteristics are shown in table 1, with similar age and sex distributions in the two cohorts. 533 CRC diagnoses (1.5%) were made within one year of FIT in the derivation cohort, and 214 (1.1%) colorectal cancer (or CRC) diagnoses within the validation cohort. The median time to diagnosis for those with a FIT<10 μg Hb in our derivation cohort with 2 years follow up was 236 (IQR 64-728) days compared to 37 (IQR 25-67) days in those with a FIT over 10 μg Hb, and 533/599 (89%) cancers were diagnosed within a year of the FIT test.

**Table 1.** Characteristics of Derivation and Validation Cohort.

| Derivation: FITs 1st November 2016- 30th November 2021 |  |  |  |  | Validation: FITs 1st December 2021 – 30<sup>th</sup> November 30th 2022 |  |  |  |
| --- | --- | --- | --- | --- | --- | --- | --- | --- |
| Test | N of patients on day of FIT | N of FITs (or patients) which had blood results 14 days after or 1 year prior to FIT | % missing | N (%) or median value (IQR) | N of patients on day of FIT | N of FITs (or patients) which had blood results 14 days after or 1 year prior to FIT | % missing | N (%) or median value (IQR) |
| Male | 15061 | 16274 | 0 | 16274 (44%) | 9227 | 8899 | 0 | 8899 (44%) |
| Female | 19374 | 20942 | 0 | 20942 (56%) | 11785 | 11335 | 0 | 11335 (56%) |
| Age[18,40] | 2247 | 2278 | 0 | 2278 (6%) | 2373 | 2350 | 0 | 2350 (12%) |
| Age[40,55] | 7210 | 7516 | 0 | 7516 (20%) | 4879 | 4753 | 0 | 4753 (23%) |
| Age[55,70] | 10991 | 11738 | 0 | 11738 (32%) | 6208 | 5962 | 0 | 5962 (29%) |
| Age[70,85] | 11803 | 12927 | 0 | 12927 (35%) | 6181 | 5859 | 0 | 5859 (29%) |
| Age[85,120] | 2598 | 2757 | 0 | 2757 (7%) | 1378 | 1310 | 0 | 1310 (6%) |
| FIT | 34435 | 37216 | 0 | 4 (4,8) | 21012 | 20234 | 0 | 4 (4,10) |
| Hb | 7666 | 33694 | 9.5 | 131 (118,143) | 631 | 17678 | 12.6 | 134 (121,145) |
| MCV | 962 | 33618 | 9.7 | 92.1 (88.1,95.9) | 631 | 17677 | 12.6 | 92.5 (88.6,96) |
| PLATELET | 962 | 33586 | 9.8 | 268 (223,322) | 627 | 17660 | 12.7 | 268 (224,321) |
| FERRITIN | 970 | 30725 | 17.4 | 65 (23,139) | 639 | 16073 | 20.6 | 74 (31,152) |
| IRON | 30 | 2956 | 92.1 | 9.1 (5.9,14.2) | 37 | 2030 | 90 | 10.7 (6,16) |
| TRANSAT | 18 | 2338 | 93.7 | 16 (10,26) | 29 | 1722 | 91.5 | 19 (10,29) |
| FCP | 6897 | 7669 | 79.4 | 30 (13,85) | 5857 | 5565 | 72.5 | 19 (5.8,46) |
| Colorectal cancer | 533 | 0 | 0 | 1.5 | 214 | 0 | 0 | 1.1 |
| Colorectal cancer death | 79 | 0 | 0 | 0.2 | 19 | 0 | 0 | 0.1 |
| Non colorectal cancer death | 1469 | 0 | 0 | 4.3 | 684 | 0 | 0 | 3.4 |

After taking the first FIT from both cohorts to calculate the performance characteristics for the validation analysis, there were 34,231 patients in the derivation cohort with 516 cancers, and 16,735 patients in the validation cohort with 206 cancers.

Table 1 summarises the cohorts, with missing haemoglobin (9.5%), MCV (9.7%), and platelet count (9.8%) blood values within the year prior and 14 days post FIT in the derivation cohort, increasing to 13% for the validation cohort (table 1). Measures of ferritin, iron, transferrin saturation and faecal calprotectin were missing in 20-90% of patients and were therefore not used in model building. There was adequate mixing of the imputed values of haemoglobin, MCV, and platelet count after the first couple of iterations within both the derivation and validation cohort (supplementary figures S1 & S2).

### Predictor Model building

#### Cox Proportional Hazards Survival Model

In the survival Cox model multiple fractional polynomial transformations were selected for age, f-Hb, and platelet count (figures S3-S5), with MCV included as a linear variable and sex as a binary variable. Haemoglobin was not selected. The fitted model is shown in table S1, with the fitted equation predicting one-year survival from CRC in table S2. There was minimal evidence for interactions between the transformed covariates (all p > 0.1, generalised likelihood ratio tests, table S3). The concordance for the model in the derivation cohort pooled between the imputed datasets was C = 0.937 (0.916-0.957).

### Performance in the validation cohort

#### Cox Proportional Hazards Survival Model

##### Stratified C-statistic performance

Concordance, as measured by a pooled C-statistic, remained highest in younger patients across the derivation and validation cohorts, with no clear drop in performance in the validation cohort (table 2). Similarly, the performance did not drop within the ethnicities, although there was greater uncertainty in the smaller strata for ethnicities other than White.

**Table 2.** Stratified C-statistics calculated for the Cox proportional hazards model with age, sex and blood tests and pooled across 10 imputed datasets in the derivation and validation cohorts (first FIT per patient only)

|  | Derivation: FITs 1st November 2016- 30th November 2021 |  |  | Validation: FITs 1st December 2021 – May 31<sup>st</sup> 2022 |  |  |
| --- | --- | --- | --- | --- | --- | --- |
| Strata | N | C-statistic | 95% CI | N | C-statistic | 95% CI |
| 18-50 years | 6093 | 0.95 | 0.91 to 1 | 4742 | 0.94 | 0.87 to 1.0 |
| 51-70 years | 14066 | 0.94 | 0.92 to 0.97 | 6634 | 0.92 | 0.89 to 0.95 |
| 71-80 years | 8348 | 0.91 | 0.89 to 0.94 | 3257 | 0.93 | 0.9 to 0.96 |
| >80 years | 5724 | 0.89 | 0.86 to 0.91 | 2102 | 0.91 | 0.88 to 0.95 |
| White | 24223 | 0.94 | 0.92 to 0.96 | 10903 | 0.93 | 0.9 to 0.95 |
| Asian | 1458 | 0.93 | 0.87 to 0.99 | 783 | 0.96 | 0.92 to 0.99 |
| Black | 853 | 0.9 | 0.79 to 1 | 476 | 0.94 | 0.87 to 1.00 |
| Other | 658 | 0.9 | 0.79 to 1 | 405 | 0.92 | 0.76 to 1.00 |
| Not recorded | 7039 | 0.92 | 0.88 to 0.95 | 4168 | 0.94 | 0.92 to 0.97 |

##### Calibration

Figure S6 shows that although there was some reduction in the calibration of the Cox model in the validation cohort compared to the derivation it remained acceptable and did not need recalibrating, allowing for the increased variability from smaller numbers.

##### Performance

A f-Hb cut-off of 10 or greater was equivalent to a 1-year cancer risk of 0.64% in the derivation cohort. We combined this threshold with the Cox model predicted cancer risk thresholds of 1%, 2% and 3% to calculate the positive predictive value, negative predictive value, sensitivity and specificity of the Cox model compared to a f-Hb ≥10 and f-Hb ≥40 cut off in the derivation and validation cohort (table 3). The Cox model with blood tests had an absolute 1% to 2% increase in the positive predictive value with a similar negative predictive value compared to a binary f-Hb cut off. This was reflected in the improvement in specificity at the expense of sensitivity using the Cox model with blood tests compared to binary f-Hb cut offs and a Cox model with only f-Hb, age and sex using similar fractional polynomial transformations to those in the full Cox model (supplementary table S6).

**Table 3.** Positive predictive value, negative predictive value, sensitivity, and specificity, at different thresholds for predicted colorectal cancer free risk using one year Kaplan Meier estimates. Cox model performance using multiple fractional polynomial transformations can be compared to equivalent binary FIT cut off rows, eg. 0,64% to f-Hb 10 and 3% to 40. (First FIT per patient only, 95% confidence intervals derived through bootstrapping (n=1000))

| Selected cut off for referral for further investigations |  | Derivation: FITs 1st November 2016- 30th November 2021 |  |  |  | Validation: FITs 1st December 2021 - 30th November 2022 |  |  |  |
| --- | --- | --- | --- | --- | --- | --- | --- | --- | --- |
| Cancer risk threshold (1 – survival predicted from model) | Equivalent FIT only threshold approximated by linear interpolation | Positive Predictive Value from Cox model | Negative Predictive Value from Cox model | Sensitivity of Cox model | Specificity of Cox model | Positive Predictive Value from Cox model | Negative Predictive Value from Cox model | Sensitivity of Cox model | Specificity of Cox model |
| Developed model with blood tests, f-Hb, age and sex |  |  |  |  |  |  |  |  |  |
| 0.64% | f-Hb=10 µg/g | 0.0649<br>(0.0593 to 0.0706) | 0.9983<br>(0.9978 to 0.9988) | 0.9163<br>(0.8902 to 0.9404) | 0.797<br>(0.7924 to 0.8014) | 0.0611<br>(0.0325 to 0.043) | 0.999<br>(0.998 to 0.999) | 0.923<br>(0.89 to 0.962) | 0.822<br>(0.696 to 0.71) |
| 1% | f-Hb=13 µg/g | 0.0792<br>(0.0723 to 0.0862) | 0.9979<br>(0.9973 to 0.9984) | 0.8906<br>(0.8619 to 0.9181) | 0.8408<br>(0.8368 to 0.8447) | 0.0759<br>(0.0325 to 0.043) | 0.999<br>(0.998 to 0.999) | 0.912<br>(0.89 to 0.962) | 0.861<br>(0.696 to 0.71) |
| 2% | f-Hb=28 µg/g | 0.1032<br>(0.094 to 0.1125) | 0.9973<br>(0.9967 to 0.9979) | 0.8493<br>(0.8165 to 0.8816) | 0.8864<br>(0.8829 to 0.89) | 0.0976<br>(0.0325 to 0.043) | 0.998<br>(0.998 to 0.999) | 0.874<br>(0.89 to 0.962) | 0.899<br>(0.696 to 0.71) |
| 3% | f-Hb=40 µg/g | 0.1239<br>(0.1127 to 0.1352) | 0.9968<br>(0.9961 to 0.9975) | 0.8178<br>(0.7804 to 0.8523) | 0.911<br>(0.9078 to 0.9143) | 0.115<br>(0.0325 to 0.043) | 0.998<br>(0.998 to 0.999) | 0.83 (0.89 to 0.962) | 0.92<br>(0.696 to 0.71) |
| Developed model with f-Hb, age and sex |  |  |  |  |  |  |  |  |  |
| 0.64% | f-Hb=10 µg/g | 0.0629<br>(0.0574 to 0.0684) | 0.9982<br>(0.9976 to 0.9986) | 0.9099<br>(0.8834 to 0.9343) | 0.7914<br>(0.7871 to 0.7957) | 0.0461<br>(0.0398 to 0.0525) | 0.999<br>(0.998 to 0.999) | 0.931<br>(0.892 to 0.964) | 0.759<br>(0.752 to 0.765) |
| 1% | f-Hb=13 µg/g | 0.0756<br>(0.069 to 0.0824) | 0.9977<br>(0.9971 to 0.9982) | 0.8812<br>(0.852 to 0.9085) | 0.8343<br>(0.8303 to 0.8382) | 0.0689<br>(0.0596 to 0.0783) | 0.999<br>(0.998 to 0.999) | 0.924<br>(0.884 to 0.959) | 0.844<br>(0.838 to 0.849) |
| 2% | f-Hb=28 µg/g | 0.0968<br>(0.0882 to 0.1056) | 0.9971<br>(0.9965 to 0.9977) | 0.841<br>(0.8087 to 0.8727) | 0.8792<br>(0.8757 to 0.8827) | 0.0854<br>(0.0736 to 0.0972) | 0.998<br>(0.998 to 0.999) | 0.891<br>(0.844 to 0.934) | 0.88<br>(0.875 to 0.885) |
| 3% | f-Hb=40 µg/g | 0.1141<br>(0.1037 to 0.1245) | 0.9965<br>(0.9959 to 0.9972) | 0.8008<br>(0.7659 to 0.8358) | 0.9043<br>(0.9012 to 0.9075) | 0.0967<br>(0.0831 to 0.11) | 0.998<br>(0.997 to 0.999) | 0.857<br>(0.806 to 0.906) | 0.9 (0.895 to 0.904) |
| Developed model with f-Hb only |  |  |  |  |  |  |  |  |  |
| f-Hb ≥ 10 µg/g |  | 0.0607<br>(0.0554 to 0.0661) | 0.998<br>(0.9974 to 0.9985) | 0.8988<br>(0.8712 to 0.9244) | 0.7861<br>(0.7818 to 0.7905) | 0.0492<br>(0.0425 to 0.0559) | 0.9988<br>(0.9982 to 0.9994) | 0.9305<br>(0.8916 to 0.9639) | 0.7747<br>(0.7682 to 0.7809) |
| f-Hb ≥ 40 µg/g |  | 0.1172<br>(0.1065 to 0.128) | 0.9962<br>(0.9955 to 0.9969) | 0.778<br>(0.741 to 0.8136) | 0.9098<br>(0.9067 to 0.9129) | 0.0971<br>(0.0835 to 0.111) | 0.998<br>(0.997 to 0.998) | 0.837<br>(0.783 to 0.888) | 0.903<br>(0.898 to 0.907) |

##### Net benefit analysis

Figure 1 shows the net benefit plots for the derivation (figure 1a) and validation (figure 1b) Cox models comparing the balance between true positives and false positives, weighted for the different cancer threshold probabilities that can trigger referral to secondary care. This shows that at all thresholds there was a net benefit using the Cox model with blood tests compared to f-Hb only. Extrapolating true and false positive and negative rates to 100,000 FITs in the validation cohort showed that using the Cox model with blood tests reduced the number of normal colonoscopies needed by 1,729 colonoscopy tests (95% CI 1458 to 2007) compared to ≥ 40 f-Hb cut off, and to 4,716 colonoscopy tests (95% CI 4,257 to 5,177) compared to ≥ 10 f-Hb cut off, an 18-40% decrease. There was no significant predicted change in missed cancers (+8 compared to ≥ 40 f-Hb cut off (95% CI –43 to 63) and +9 compared to ≥ 10 f-Hb cut off (95% CI –3 to 29). When compared to the Cox model with only f-Hb, age, and sex, the addition of blood tests similarly reduced the number of colonoscopies that would be required (Table 4).

**Figure 1.**
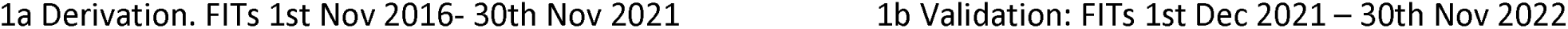
(a and b). Net benefit plots comparing the developed Cox model against f-Hb only models at different cancer risk referral thresholds using Kaplan Meier estimates. First FIT per patient only.

**Table 4.** Extrapolating true and false positive and negative rates from Cox model to 100,000 FITs in the validation cohort. (First FIT per patient only, 95% confidence intervals derived through bootstrapping (n=1000)). All numbers rounded to the nearest integer.

|  | Derivation: FITs 1st November 2016- 30th November 2021 |  |  |  | Validation: FITs 1st December 2021– 30th November 2022 |  |  |  |
| --- | --- | --- | --- | --- | --- | --- | --- | --- |
| Cancer risk threshold | Colonoscopies performed for patients above predicted threshold | Detected cancer cases (True positives) | Missed cancer cases (False negatives) | Normal colonoscopies (False positives) | Colonoscopies performed for patients above predicted threshold | Detected cancer cases (True positives) | Missed cancer cases (False negatives) | Normal colonoscopies (False positives) |
| Developed model with blood tests, f-Hb, age and sex |  |  |  |  |  |  |  |  |
| 0% (refer everyone) | 100000 | 1515 (1386 to 1643) | 0 (0 to 0) | 98485 (98357 to 98614) | 100000 | 1237 (1070 to 1404) | 0 (0 to 0) | 98763 (98596 to 98930) |
| 0.6% | 21383 (20928 to 21837) | 1388 (1265 to 1510) | 127 (89 to 168) | 19994 (19557 to 20441) | 18681 (18064 to 19319) | 1142 (983 to 1305) | 95 (49 to 149) | 17539 (16940 to 18158) |
| 1% | 17033 (16634 to 17434) | 1349 (1225 to 1471) | 166 (123 to 212) | 15684 (15298 to 16071) | 14856 (14305 to 15435) | 1128 (971 to 1290) | 109 (60 to 166) | 13728 (13199 to 14285) |
| 2% | 12472 (12100 to 12833) | 1287 (1168 to 1407) | 228 (176 to 281) | 11185 (10830 to 11532) | 11080 (10589 to 11586) | 1081 (925 to 1241) | 155 (95 to 225) | 9998 (9533 to 10479) |
| 3% | 10001 (9664 to 10336) | 1239 (1122 to 1355) | 276 (219 to 337) | 8762 (8444 to 9077) | 8917 (8479 to 9358) | 1027 (877 to 1183) | 210 (141 to 289) | 7890 (7471 to 8315) |
| Developed model with f-Hb, age and sex |  |  |  |  |  |  |  |  |
| 0% (refer everyone) | 100000 | 1515 (1386 to 1643) | 0 (0 to 0) | 98485 (98357 to 98614) | 100000 | 1237 (1070 to 1404) | 0 (0 to 0) | 98763 (98596 to 98930) |
| 0.6% | 21919 (21481 to 22357) | 1379 (1255 to 1501) | 136 (98 to 179) | 20540 (20117 to 20968) | 24954 (24314 to 25605) | 1151 (993 to 1315) | 85 (43 to 134) | 23802 (23165 to 24453) |
| 1% | 17657 (17256 to 18057) | 1335 (1216 to 1457) | 180 (136 to 228) | 16321 (15935 to 16716) | 16594 (16044 to 17174) | 1143 (984 to 1306) | 94 (49 to 147) | 15451 (14913 to 16013) |
| 2% | 13169 (12807 to 13523) | 1274 (1157 to 1393) | 241 (189 to 295) | 11895 (11553 to 12240) | 12907 (12411 to 13427) | 1102 (946 to 1265) | 135 (80 to 196) | 11805 (11330 to 12310) |
| 3% | 10637 (10309 to 10964) | 1213 (1098 to 1330) | 302 (242 to 362) | 9423 (9114 to 9735) | 10971 (10511 to 11461) | 1061 (907 to 1217) | 176 (113 to 244) | 9910 (9470 to 10385) |
| Developed model f-Hb only |  |  |  |  |  |  |  |  |
| f-Hb $\geq 10$ | 22427 (21989 to 22865) | 1362 (1241 to 1484) | 153 (113 to 197) | 21065 (20641 to 21490) | 23406 (22767 to 24052) | 1151 (992 to 1314) | 86 (43 to 135) | 22255 (21634 to 22893) |
| f-Hb $\geq 40$ | 10061 (9740 to 10377) | 1179 (1065 to 1294) | 336 (275 to 399) | 8882 (8584 to 9184) | 10654 (10206 to 11132) | 1035 (883 to 1190) | 202 (135 to 276) | 9619 (9186 to 10079) |

These illustrative numbers for predicted outcomes were calculated by multiplying the true and false, positive and negative rates 100,000. These rates were calculated using Kaplan Meier estimates above and below each threshold level following the approach described by Vickers et al<sup>19</sup>.

### Performance in the ELHT cohort

#### Cox Proportional Hazards Survival Model

In total 30291 patients had FITs undertaken in primary care with symptoms of suspected CRC from 2019-2023 with 328 (1.1%) CRC identified in the year following the test. The proportion of those with a FIT above the threshold of 10 µg/g was 21.8% (6616/30291). Of the CRCs diagnosed 295 were above the threshold of 10 µg/g whilst 287 were above the predicted 1% one-year risk of CRC therefore 8 additional cancers would be missed using the 1% predicted risk. However, 6288 tests were above the threshold of 10 µg/g for referral whilst only 4793 were above the 1% predicted risk threshold therefore potentially saving 1495 colonoscopies. This would represent an additional 26 missed cancers per 100,000 FITs performed set against a reduction in colonoscopies of 4935 in this referral population.

## Discussion

We show that incorporating simple blood test results, age and sex with f-Hb into a clinical risk prediction model for patients with symptoms of possible CRC may improve the diagnostic pathway. Our models, which included f-Hb, age, sex, MCV and platelet count, increased the positive predictive value for CRC compared to the f-Hb cut off of ≥10 µg/g currently recommended by NICE and others, with minimal change in the negative predictive value^6^ ^21^ ^22^. The corresponding reduction in false positive rate could lead to fewer referrals and colonic investigations. During validation, the false negative rate increased resulting in more missed cancers, but to a smaller extent than the reduction in false positives. Compared to f-Hb ≥10 µg/g the Cox model, if implemented in a population of 100,000 people having FITs, might reduce the number of referrals and tests by >4700 with minimal change in the number of cancers that would be missed. Using a f-Hb ≥10 µg/g was associated with a cancer risk of 0.64% which is considerably below the current recommended urgent referral guidelines of 3%^5^. At risk thresholds up to a 5% risk of CRC our Cox model showed similar improvements over the equivalent binary f-Hb cut offs or f-Hb modelled as a continuous variable. These findings, at a risk threshold of 1%, were similar in our second external validation using data from the north-west of England, yet further external validation is needed to assess generalisability to other populations and health care settings and the value of these trade-offs requires health economic assessment^18^. This work is ongoing as part of the COLOFIT programme.

The strengths of this study are the large population-based cohort of patients with symptomatic FIT testing over a period of 6 years. All relevant routine blood tests and CRC diagnoses were recorded within electronic health records, including complete follow up for death and its cause. Our choice of modelling the covariates as continuous transformations avoids the pitfalls of selective cut offs and crude thresholds. This allows a more personalised approach to individual risk that is more informative and potentially more useful for the health care system when prioritising who to investigate in diagnostic CRC pathways. We have incorporated the effect of missing data within our models and undertaken a targeted validation within a separate cohort of more recent patients within Nottingham along with a validation in the ELHT cohort. The use of the routinely prospectively collected unselected health data minimised selection bias and increased the generalisability of the final model. There are some limitations, however. We were unable to utilise some indices for anaemia such as ferritin due to the high proportion of missing data for these individual measures. We had smaller numbers for our validation cohorts and did not include repeat tests. We plan to continue to reassess the calibration and performance of the Cox model as data accrues and validate the model in external datasets within different referral regions to assess how transferrable the model is^14^. We identified fewer cases of CRC in the validation cohort which may reflect changes in the tested population over time, such as the inclusion of those with rectal bleeding and wider use of the test which has occurred since the pandemic. Nonetheless, the population within the validation cohort represents a ‘panrisk’ group of patients where CRC is a possibility, where other factors to discriminate those at risk of CRC are needed. Despite this population change, the internal-external validation showed the performance of the Cox model did not decrease over time in the same Nottinghamshire population: indeed, performance was slightly superior in the validation cohort (and similar at the 1% threshold in the ELHT cohort). Not all patients were investigated as this algorithm was developed using real world data – and in an ideal world all patients in the study (circa 50000) would have undergone bowel imaging (with either endoscopy or CTC) to confirm the diagnostic accuracy results. However, this was not possible without introducing selection and consent into the study due to the patient risk of the investigations, and this would thereby reduce the generalisability of any results. Data on family history and symptoms are not available in the data used in this study. We have previously published on the association of symptoms and the outcome of CRC in Nottingham early in its course, where we had access to paper referrals, finding that symptoms were not associated with predicting the risk of CRC^29^. Furthermore, most patients with symptomatic CRC present with a low-risk symptom and current NICE guidance has therefore focused on the role of other methods (FIT) to identify those presenting in primary care with such symptoms at most risk of CRC^31^, hence our focus on FIT. We have modelled the risk of CRC but other diagnoses are made in patients urgently referred for possible cancer, so further work will be required to detail the potential benefits of, for example, diagnosing inflammatory bowel disease and polyps. This work is currently not possible with the dataset available but clearly needs to be considered in terms of diagnostic delay and possible malignant transformation of polyps. However FIT is currently used as a gateway test for referral for CRC rather than these other pathologies so we have focused our outcome on CRC. Further work is planned to determine how best to enact these thresholds within a 2WW pathway and how best to communicate these risks (and resource implications) to clinicians, policy makers and patients.

We also did several sensitivity analysis and comparisons between Cox and logistic modelling. When we modelled fHb as a continuous variable, transformed, and added either age or age and sex none of these models performed as well as the more complex Cox model we developed. When comparing Cox and logistic modelling techniques we found the differences were minor in terms of performance but calibration in validation was superior for the Cox model.

There have been some prior attempts to utilise blood tests to improve the performance of FIT. In a study of 16,604 FITs in a patient cohort in Oxford three models were developed combining f-Hb, age and sex. The authors found no additional benefit from the use of blood tests; however, the cohort described derived from patients fulfilling the DG30 criteria (i.e. of lower risk of cancer), FITs were analysed on the HM JACK-arc rather than the OC Sensor analyser, and, importantly, FITs were undertaken using faecal samples collected into universal stool collection pots and transported to the laboratory before being transferred in to stabilising buffer^23^. Haemoglobin degradation is likely to have occurred during this time^24^. The COLONPREDICT study included f-Hb, age, sex, rectal bleeding, benign anorectal lesions, rectal mass, serum carcinoembryonic antigen, blood haemoglobin, colonoscopy in the last 10 years, and treatment with aspirin^25^. We were unable to include family history in the model developed in Nottingham as this was not available from the data used (and is not routinely captured in the NHS). We found that MCV improved model FIT more than haemoglobin, possibly because it is a better surrogate for iron deficiency anaemia and is less affected by gender. The FAST score did not incorporate blood tests into the risk prediction model, instead using f-Hb level, age and sex^26^. A further study modelling the risk of CRC using logistic regression in a population in Scotland concluded no benefit to the use of a risk score in those with a low FIT however it suggested that modelled risk might allow the raising of the current threshold and thus reduce endoscopy demand^27^. A recent systematic review of the performance of f-Hb-based risk prediction models identified 22 studies combining FIT with one or more variable to predict the risk of CRC or advanced colonic polyps^11^. The review found that 10 studies reported development of a model, whilst four reported validation of models and three presented both derivation and validation^11^. The models were developed in modestly sized cohorts and were considered methodologically poor with a lack of validation. None presented a net-benefit analysis.

While the output of the models require a calculator for computation, the aim of the study was to identify the best fitting model which yielded net benefit, regardless of complexity. Equations are implementable within current NHS IT systems with examples of successful implementation of model-based decision making in primary care, including the QRisk algorithms^28^. The advantage of this approach is that it allows calculation of individualised risk prediction using available results and demographics such that a tailored approach could be considered i.e. we could determine at a given level of risk of CRC what f-Hb result (incorporating age, sex and FBC) should trigger referral for further investigation. Table 5 shows some clinical scenarios of how the predicted risk varies for a woman with a f-Hb of 40 µg/g depending on her full blood count results and age.

**Table 5.**
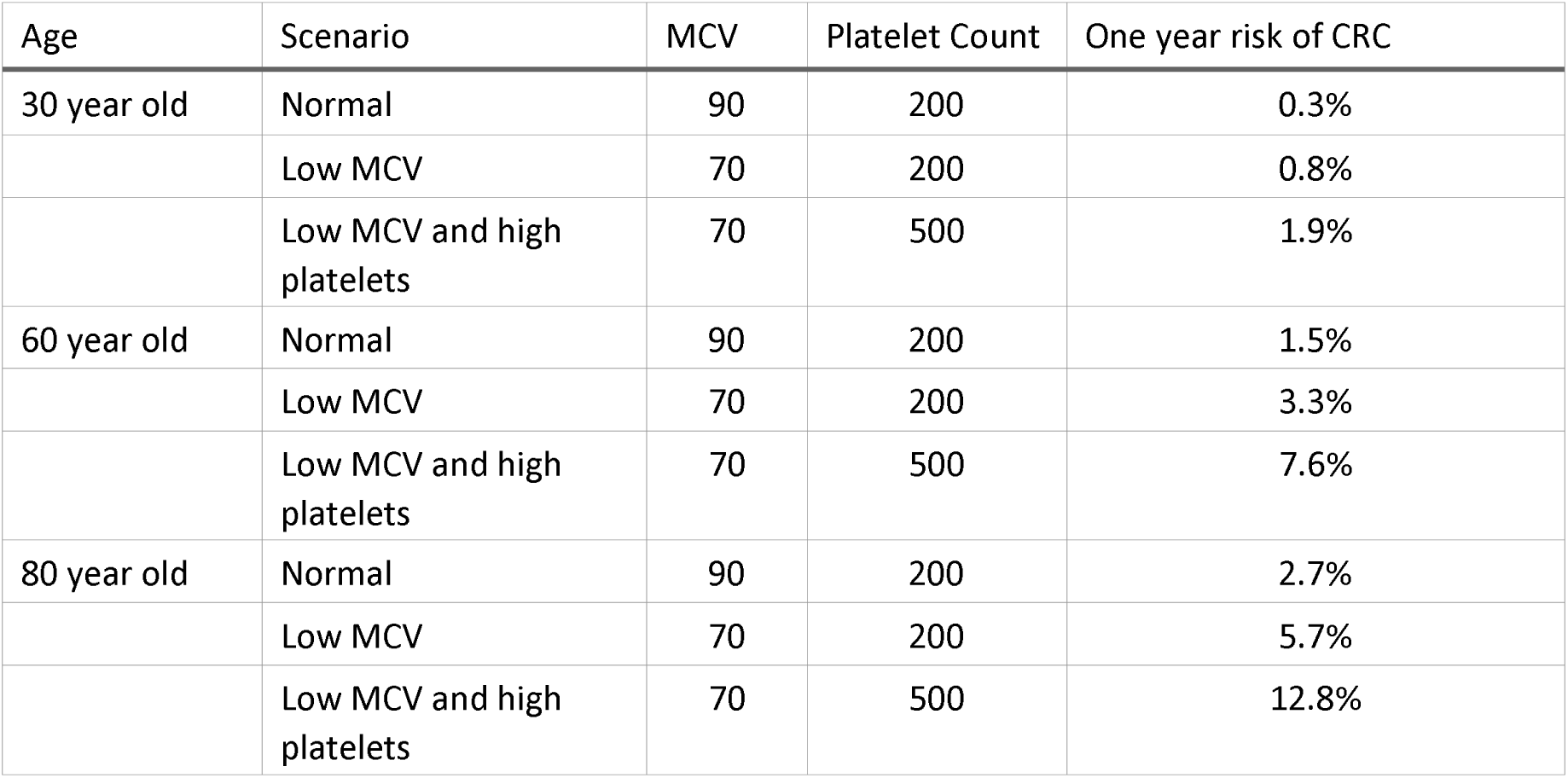
Risk prediction for illustrative clinical scenarios for a woman with a f-Hb result of 40 µg/g using a Cox model with age, sex and blood tests to predict one year risk of colorectal cancer.

| Age | Scenario | MCV | Platelet Count | One year risk of CRC |
| --- | --- | --- | --- | --- |
| 30 year old | Normal | 90 | 200 | 0.3% |
|  | Low MCV | 70 | 200 | 0.8% |
|  | Low MCV and high platelets | 70 | 500 | 1.9% |
| 60 year old | Normal | 90 | 200 | 1.5% |
|  | Low MCV | 70 | 200 | 3.3% |
|  | Low MCV and high platelets | 70 | 500 | 7.6% |
| 80 year old | Normal | 90 | 200 | 2.7% |
|  | Low MCV | 70 | 200 | 5.7% |
|  | Low MCV and high platelets | 70 | 500 | 12.8% |

Implementation of such an approach requires a re-evaluation of the guidance on prioritising investigation for CRC. At present the aspiration in the English NHS is to refer people being assessed in primary care with a risk of cancer of 3% for further rapid investigation^31^. In CRC this threshold is de facto much lower, as the current f-Hb threshold of ≥10 µg/g represents a risk of CRC of 1% or lower. Stakeholder consensus would be required to decide what the level of CRC risk should be to trigger referral and investigation. This would determine the diagnostic yield and health service burden all stakeholders are willing and able to accept and deliver. For example, if a 3% risk of CRC were to be implemented this would be equivalent to enacting a f-Hb threshold of ≥40 µg/g^10^. This would reduce the number of normal colonoscopies required substantially but at a cost of more missed cancers.

Our Cox model at both a 1% or 3% threshold of CRC risk would offer additional improvements in false negative and false positive rates compared to the equivalent f-Hb ≥10 µg/g or f-Hb ≥40 µg/g thresholds. In addition, if the level of f-Hb in the national Bowel Cancer Screening Programme (BCSP) is reduced alongside the expansion of the age inclusion criteria, a currently unfeasible increase in capacity will be required to deliver all the extra colonoscopies. Enacting model-based triage in symptomatic patients suspected of CRC should free up colonoscopy resource to allow expansion of the BCSP, pending further externally validation of our model in other populations. This approach to risk prediction incorporating demographics and blood tests could be extended to the asymptomatic screening population where there may be the possibility to improve the selection of patients for colonoscopy if the screening referral values for FIT are changed.

In conclusion, enacting a model-based triage of a symptomatic CRC pathway could decrease the burden on endoscopy whilst maintaining diagnostic accuracy as targeted validation of our Cox model suggested that using the model may lead to a similar proportion of cancers detected whilst reducing the number of colonoscopies performed compared to the equivalent binary f-Hb cut offs. The current f-Hb cut off of 10 µg/g or greater is equivalent to an individual CRC risk less than one percent, resulting in many false positives and therefore colonoscopies that is arguably unsustainable within the health system.

## Notes

### Competing Interest Statement

CJR has received grant funding from ARC medical, Norgine. Medtronic, 3D Matrix solutions and Olympus medical. He was an expert witness for ARC medical and Olympus medical

### Summary of Updates

We have updated the manuscript following peer review comments and included an external validation of the COLOFIT algorithm in data from Lancashire in the UK.

